# A two-pronged approach for rapid and high-throughput SARS-CoV-2 nucleic acid testing

**DOI:** 10.1101/2020.12.04.20234450

**Authors:** Neeru Gandotra, Irina Tikhonova, Nagarjuna R. Cheemarla, James Knight, Ellen Foxman, Antonio Giraldez, Peidong Shen, Kaya Bilguvar, Curt Scharfe

## Abstract

Improved molecular screening and diagnostic tools are needed to substantially increase SARS-CoV-2 testing capacity and throughput while reducing the time to receive test results. Here we developed multiplex reverse transcriptase polymerase chain reaction (m-RT-PCR) for detection of SARS-CoV-2 using rapid DNA electrophoresis and alternatively using multiplex viral sequencing (mVseq). For RNA specimens extracted from nasopharyngeal (NP) swabs in viral transport media (VTM), our assays achieved a sensitivity for SARS-CoV-2 detection corresponding to cycle threshold (Ct) of 37.2 based on testing of these specimens using quantitative reverse transcription PCR (RT-qPCR). For NP swab-VTM specimens without prior RNA extraction, sensitivity was reduced to Ct of 31.6, which was due to lower concentration of SARS-CoV-2 genome copies in VTM compared to RNA-extracted samples. Assay turnaround time was 60 minutes using rapid gel electrophoresis, 90 minutes using Agilent Bioanalyzer, and 24-48 hours using Illumina sequencing, the latter of which required a second PCR to produce a sequence-ready library using m-RT-PCR products as the template. Our assays can be employed for high-throughput sequencing-based detection of SARS-CoV-2 directly from a clinical specimen without RNA isolation, while ease-of-use and low cost of the electrophoresis-based readout enables screening, particularly in resource-constrained settings.

## Introduction

The emergence of worldwide Coronavirus disease 2019 (COVID-19) has spurred technology development for SARS-CoV-2 molecular diagnostic testing. Traditional viral nucleic acid testing (NAT) has relied on extracting RNA from patient samples such as nasopharyngeal (NP) swabs in viral transport media (VTM) followed by virus detection using RT-qPCR^1,2^. Scalability of this workflow during this COVID-19 pandemic has been hampered by supply shortages in NP swabs and RT-qPCR assay reagents, and by limited sample throughput capabilities and extended time to report test results. Recently, several innovative approaches for SARS-CoV-2 testing have been developed such as analysis of human saliva specimens with^3-7^ or without a prior RNA purification step^8^, a rapid colorimetric assay^9^ or a CRISPR-based assay^10^ using reverse-transcription loop-mediated isothermal amplification (RT-LAMP), an amplification-free CRISPR-Cas13a-based mobile phone assay^11^, and SARS-CoV-2 detection using next-generation sequencing as a readout^12-15^. Successful implementation of these approaches into workable testing solutions could dramatically increase SARS-CoV-2 diagnostic capacity and throughput while reducing the time to receive test results.

Here, we developed a two-pronged approach for rapid and for high-throughput SARS-CoV-2 nucleic acid testing (**Figure 1**). Our approach utilizes multiplex reverse transcriptase polymerase chain reaction (m-RT-PCR) to capture three SARS-CoV-2 genomic regions plus the human *RPP30* control gene in a single-tube reaction (4-plex) directly from a small VTM sample without prior RNA extraction. DNA gel-electrophoresis was developed for the rapid readout of m-RT-PCR amplification products based on their designed size differences. Alternatively, automated DNA fragment detection using the Agilent Bioanalyzer instrument was employed for medium-throughput applications (12-96 samples within 90 minutes). For high-throughput SARS-CoV-2 testing of potentially tens of thousands of samples, m-RT-PCR amplification products were converted into a sequence-ready library^16^ for Illumina sequencing with results available within 24-48 hours. Additional improvements of the sequencing workflow for SARS-CoV-2 screening and diagnostic testing will enable increased capacity while reducing the time to receive test results. Major challenges remain at the “*front end*” of sample acquisition and pre-processing that will need to be addressed for high volume implementation.

**Figure 1:**
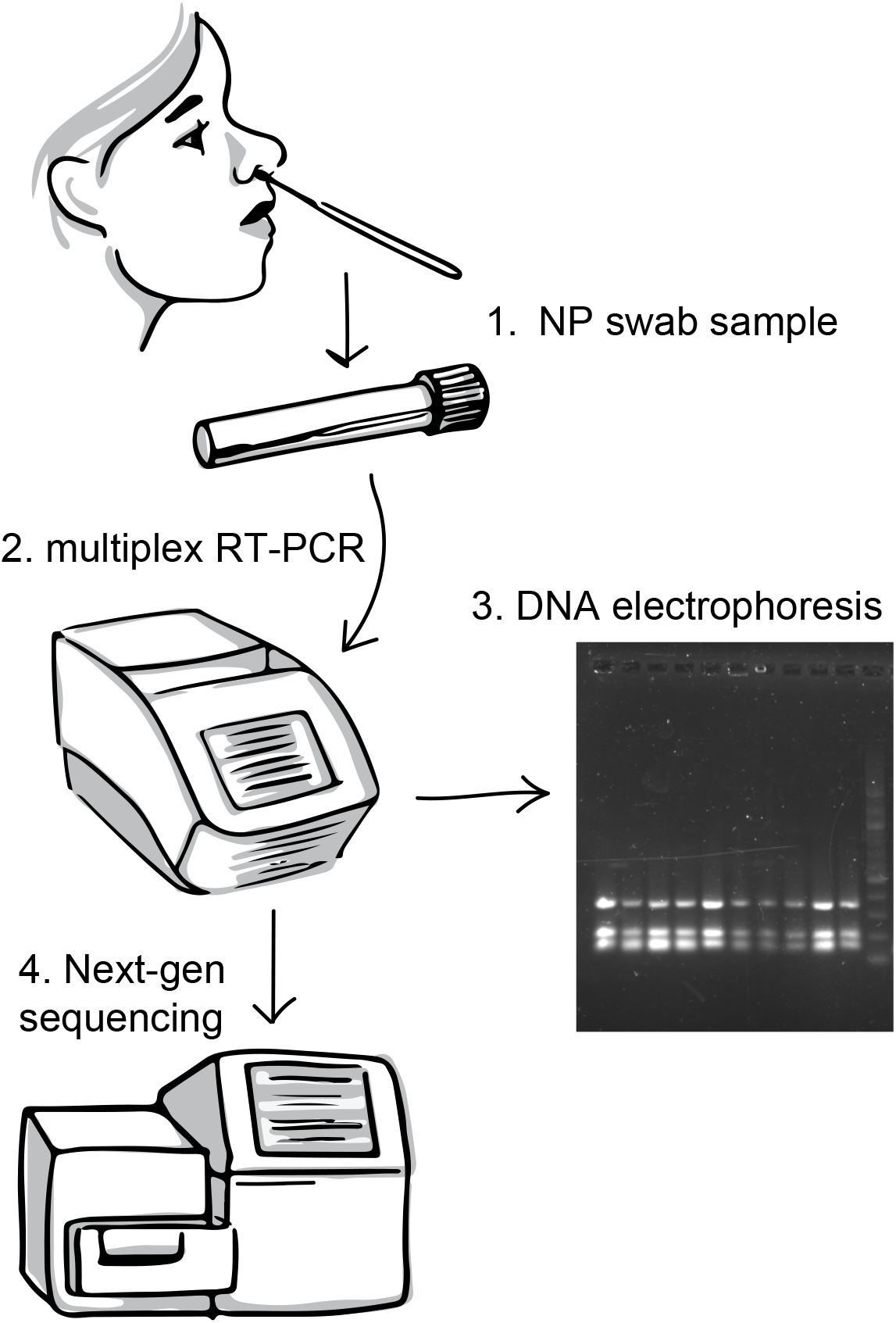
Workflow for SARS-CoV-2 detection. Overview of the assay showing the main steps of sample collection, multiplex RT-PCR, the rapid DNA electrophoresis-based readout, and alternatively the high-throughput sequencing-based readout for SARS-CoV-2 detection.

## Materials and Methods

### Study population

The Yale Institutional Review Board determined this study to be not human subjects research (IRB Protocol #2000029556). Residual nasopharyngeal (NP) swab samples from clinical testing (n=30) were obtained from the Yale-New Haven Hospital (YNHH) Clinical Virology Laboratory. Samples were de-identified and labeled only with the Ct values from clinical virology testing using RT-qPCR. NP swabs had initially been collected and placed in viral transport media (VTM) by the healthcare provider, and residual VTM was stored at −80C immediately following sample accessioning. Clinical virology testing was previously performed by extracting total nucleic acids using the NUCLISENS easyMAG platform (BioMérieux, France), followed by reverse transcriptase quantitative PCR for N1, N2, and RNAse P targets following the CDC protocol^1^. The 30 samples included 20 positive samples with higher (Ct value for N1, 12.1-24.9) and lower (Ct value for N1, 31.6-37.2) viral loads and 10 negative samples (**Table 1**).

**Table 1.** Samples studied and assay performance comparison.

| Sample | RT-qPCR testing | Ct value N1 | Sample type | Presence of SARS-CoV-2 VTM as input |  | Presence of SARS-CoV-2 RNA as input |  |
| --- | --- | --- | --- | --- | --- | --- | --- |
|  |  |  |  | Gel / Bioanalyzer | NGS | Gel / Bioanalyzer | NGS |
| S01 | SARS-CoV-2 (COVID-19) N1 RT-PCR | 20.1 | NP | Yes/Yes | Yes | NT/Yes | Yes |
| S02 | SARS-CoV-2 (COVID-19) N1 RT-PCR | 24.9 | NP | Yes/Yes | Yes | NT | NT |
| S03 | SARS-CoV-2 (COVID-19) N1 RT-PCR | 24.5 | NP | Yes/Yes | Yes | NT | NT |
| S04 | SARS-CoV-2 (COVID-19) N1 RT-PCR | 24 | NP | Yes/Yes | Yes | NT | NT |
| S05 | SARS-CoV-2 (COVID-19) N1 RT-PCR | 15.5 | NP | Yes/Yes | Yes | NT | NT |
| S06 | SARS-CoV-2 (COVID-19) N1 RT-PCR | 22.1 | NP | Yes/Yes | Yes | NT | NT |
| S07 | SARS-CoV-2 (COVID-19) N1 RT-PCR | 22.5 | NP | Yes/Yes | Yes | NT | NT |
| S08 | SARS-CoV-2 (COVID-19) N1 RT-PCR | 19.4 | NP | Yes/Yes | Yes | NT | NT |
| S09 | SARS-CoV-2 (COVID-19) N1 RT-PCR | 19.1 | NP | Yes/Yes | Yes | NT | NT |
| S10 | SARS-CoV-2 (COVID-19) N1 RT-PCR | 12.1 | NP | Yes/Yes | Yes | NT | NT |
| S11 | SARS-CoV-2 (COVID-19) N1 RT-PCR | 32.4 | NP | No/No | No | NT/Yes | Yes |
| S12 | SARS-CoV-2 (COVID-19) N1 RT-PCR | 37.2 | NP | No/No | No | NT/Yes | Yes |
| S13 | SARS-CoV-2 (COVID-19) N1 RT-PCR | 35.3 | NP | No/No | No | NT/Yes | Yes |
| S14 | SARS-CoV-2 (COVID-19) N1 RT-PCR | 35.8 | NP | No/No | No | NT/Yes | Yes |
| S15 | SARS-CoV-2 (COVID-19) N1 RT-PCR | 31.6 | NP | No/Yes | Yes | NT/Yes | Yes |
| S16 | SARS-CoV-2 (COVID-19) N1 RT-PCR | 36 | NP | No/No | No | NT/Yes | Yes |
| S17 | SARS-CoV-2 (COVID-19) N1 RT-PCR | 36.3 | NP | No/No | No | NT/Yes | Yes |
| S18 | SARS-CoV-2 (COVID-19) N1 RT-PCR | 34.7 | NP | No/No | No | NT/Yes | Yes |
| S19 | SARS-CoV-2 (COVID-19) N1 RT-PCR | 34.4 | NP | No/No | No | NT | NT |
| S20 | SARS-CoV-2 (COVID-19) N1 RT-PCR | 35.4 | NP | No/No | No | NT | NT |
| S21 | SARS-CoV-2 (COVID-19) N1 RT-PCR | 45 | NP | No/No | No | NT | NT |
| S22 | SARS-CoV-2 (COVID-19) N1 RT-PCR | 45 | NP | No/No | No | NT | NT |
| S23 | SARS-CoV-2 (COVID-19) N1 RT-PCR | 45 | NP | No/No | No | NT | NT |
| S24 | SARS-CoV-2 (COVID-19) N1 RT-PCR | 45 | NP | No/No | No | NT | NT |
| S25 | SARS-CoV-2 (COVID-19) N1 RT-PCR | 0* | NP | No/No | No | NT | NT |
| S26 | SARS-CoV-2 (COVID-19) N1 RT-PCR | 0* | NP | No/No | No | NT | NT |
| S27 | SARS-CoV-2 (COVID-19) N1 RT-PCR | 0* | NP | No/No | No | NT | NT |
| S28 | SARS-CoV-2 (COVID-19) N1 RT-PCR | 0* | NP | No/No | No | NT | NT |
| S29 | SARS-CoV-2 (COVID-19) N1 RT-PCR | 0* | OP | No/No | No | NT | NT |
| S30 | SARS-CoV-2 (COVID-19) N1 RT-PCR | 0* | NP | No/No | No | NT | NT |

The 30 samples analyzed included 10 high-positives (Ct 12.1-24.9), 10 low-positives (Ct 31.6-37.2) and 10 negative controls. Abbreviations: NP, nasopharyngeal swab; OP, oropharyngeal; NT, not tested. \*No Ct values were obtained for Cepheid only tested samples.

### Sample preparation

We adopted the protocol described by Ladha et al.^17^ to remove the initial RNA extraction step and enable direct use of NP swab-VTM specimens for SARS-CoV-2 detection. Equal volumes of VTM sample and QE Buffer (Lucigen, Cat No. QE09050) were mixed and heated at 95 °C for 5 min for sample denaturation and to release the viral RNA followed by an immediate cool down at 4 °C. A volume of 10.99 μl of the VTM-QE mix was used in the m-RT-PCR (total volume 25 μl). The single-tube multiplex RT-PCR was developed to capture three regions of the SARS-CoV-2 genome and one region of the human *RPP30* gene used to control for sample collection (**Table 2**). The oligonucleotide primers included Cov-S (22340-22447 bp) described by Bloom et al.^12^, Cov-JN described by Shirato et al.^18^ (29125-29282 bp), as well as Cov-310 (13-322 bp) and a *RPP30* amplicon (23-115 bp) identified in this study. We optimized the m-RT-PCR by testing different RT enzymes, of which the Superscript IV One step RT PCR system (Thermo Fisher Scientific, Cat No. 12594025) was identified and run using the following protocol: 48 °C for 10 min, 95 °C for 2 min, followed by 10 cycles of 95 °C for 3 s, 58 °C for 5 s, and 72 °C for 18 s, and followed by 35 cycles for 95 °C for 3 s, 72 °C for 20 s and a final extension at 72 °C for 1 minute.

**Table 2.**
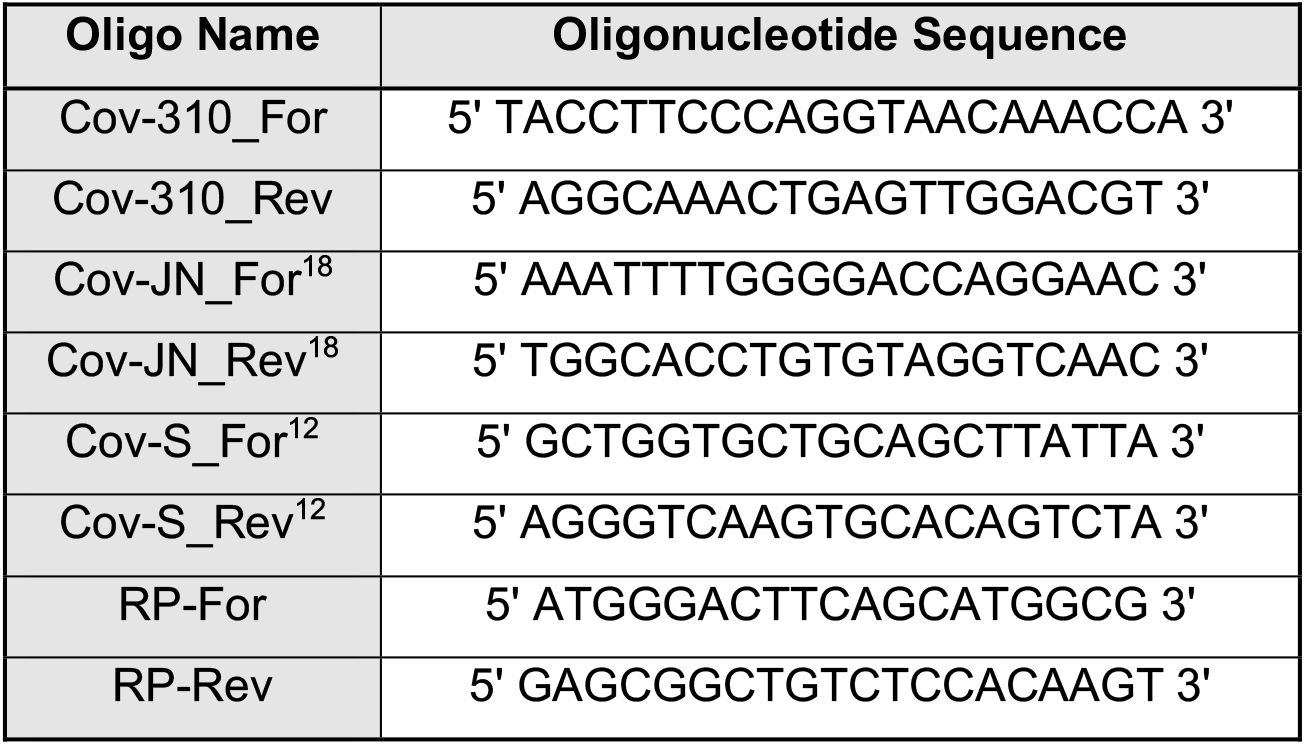
Oligonucleotide primers included in the multiplex RT-PCR.

### Detection of m-RT-PCR amplification products

The four m-RT-PCR amplicons were selected to have different lengths for size separation and analysis using DNA electrophoresis. Volumes of 5 μl or 1 μl of the m-RT-PCR product was run directly on a 1.5% agarose gel or the Agilent Bioanalyzer, respectively (**Figure 2**). Alternatively, m-RT-PCR amplicon readout was developed for next-generation sequencing (**Figure 3**). Producing the sequence-ready library containing the Illumina indexes required a second PCR using the first m-RT-PCR products as the template^16^. This second PCR was run using this protocol: 98 °C for 50 s, 14 cycles of 98 °C for 16 s, 72 °C for 20 s followed by a final extension of 72 °C for 2 min. A 10 μl aliquot of the 2^nd^ PCR amplification products from each sample (n=30) was pooled and bead-purified using a 1.8:1 ratio of Ampure XP beads (Beckman Coulter) to DNA. Following quantification, a small aliquot of the library (18 μl of 1.3 nM final concentration) was run on Illumina NovaSeq 6000 or MiSeq using paired-end 150 bp sequencing. The paired-end reads were aligned using BWA MEM to a reference containing the SARS-CoV-2 genome (NC_045512.2) and *RPP30* gene (NM_001104546.1). An in-house script parsed the alignments, and, for each pair, computed the alignments’ starting locations and strands for the pair’s two reads. Read pairs with unique alignments within 4bp of the expected amplicon endpoints, with proper alignment directionality, were counted as read pairs for that amplicon. Other reads were filtered from the counts.

**Figure 2:**
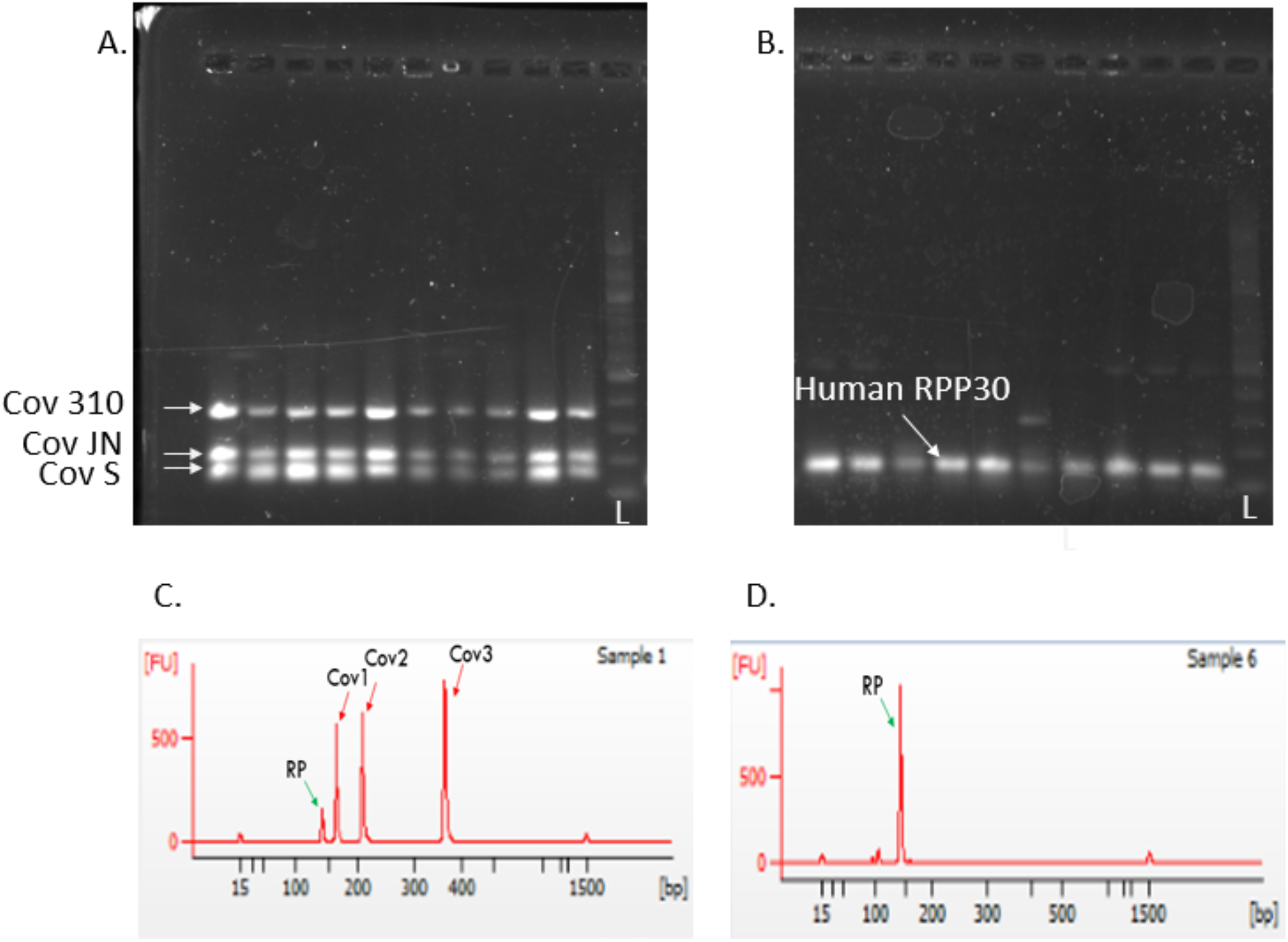
Rapid detection of SARS-CoV-2. Readout of multiplex RT-PCR amplicons was performed within 60 minutes using gel electrophoresis for (**A**) high viral load positive samples (ranging from Ct 12.1-24.9) and (**B**) SARS-CoV-2 negative specimens, and within 90 minutes using the Agilent Bioanalyzer for (**C**) high viral load samples (S01) and (**D**) negative samples (S26).

**Figure 3:**
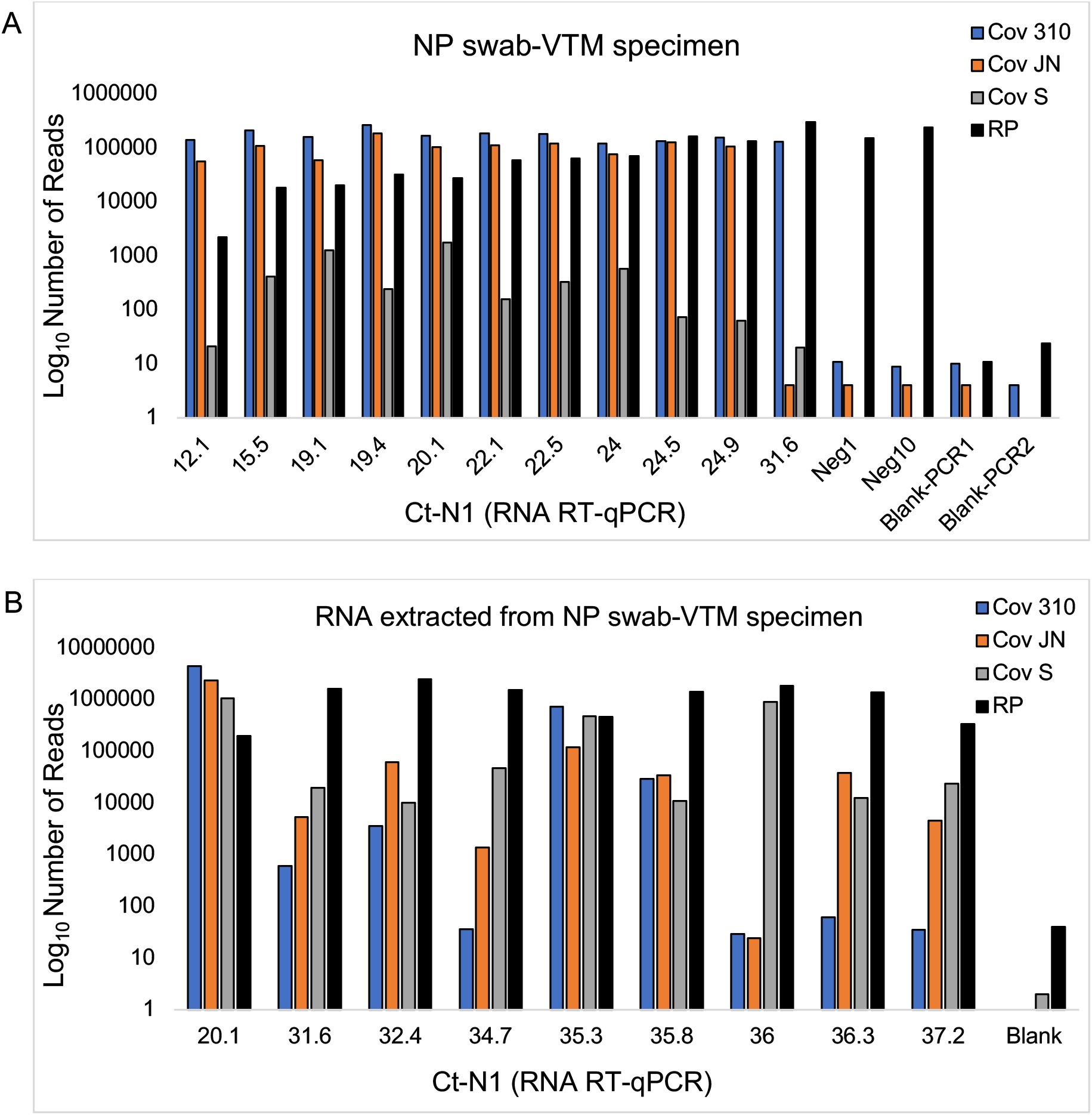
High-throughput detection of SARS-CoV-2. Readout of multiplex RT-PCR amplicons using next-generation sequencing for (**A**) NP swab-VTM specimens without prior RNA extraction, and (**B**) RNA extracted from NP specimens. Y-axis shows the log_10_ transformed sequencing reads mapping to the four amplicon targets, and X-axis shows N1 cycle thresholds (Ct) for each sample from RT-qPCR testing.

## Results

A single-tube multiplex RT-PCR (4-plex) was developed to capture three regions of the SARS-CoV-2 genome and the Human *RPP30* control amplicon from NP swab-VTM specimens. The assay was first evaluated for NP swab-VTM specimens without prior RNA extraction. Two alternative approaches for readout of m-RT-PCR amplicons were developed (**Figure 1**).

First, a fast readout was designed to return results in less than 60 minutes by analyzing the m-RT-PCR amplification products directly using agarose gel electrophoresis. SARS-CoV-2 positive VTM samples showed three bands corresponding to the three viral amplicons (**Figure 2A**), while negative samples only showed amplification of the single *RPP30* target (**Figure 2B**). The reagent and supplies cost of the gel-based assay including QE buffer, PCR reagents, primers, tubes, pipet tips, gloves, and gel was estimated at $ 5.37 per sample. Alternatively, m-RT-PCR amplification products could also be analyzed using the Agilent Bioanalyzer within 90 minutes, and with an ability to multiplex larger samples batches (e.g., 96 samples using the Agilent TapeStation, $ 1.69 per sample) at an estimated reagent and supplies cost of $ 7.01 per sample. Using the Bioanalyzer, SARS-CoV-2 positive samples showed four peaks including the human *RPP30* amplicon at a size of 145 bp, and the three viral amplicons of 157, 210 and 348 bp (**Figure 2C**). Negative samples showed only a single peak for *RPP30* (**Figure 2D**). For some samples, additional smaller peaks were seen indicating non-specific amplification products or primer dimers, which however differed in size compared to the intended targets and could thus be readily identified. We found that all NP swab-VTM specimens with higher viral loads (Ct N1, 12.1-24.9) consistently showed the three SARS-CoV-2 bands on the agarose gel and four peaks on the Bioanalyzer. Among the samples with lower viral loads (Ct N1, 31.6-37.2), only one sample (S15, Ct N1, 31.6) showed a single SARS-CoV-2 and the *RPP30* peak on the Bioanalyzer (**Supplemental Figure 1**) but not on the agarose gel. The other nine low viral-load samples did not show visible amplification products using either detection method.

Second, high-throughput readout of m-RT-PCR products using next-generation sequencing was developed (**Figure 3A**). Similar to results above, we found that VTM samples with high viral loads consistently generated a sufficient number of reads for at least one SARS-CoV-2 amplicon and for *RPP30*, while samples with lower viral load did not show read counts that were significantly different from the small level of background reads in negative control samples. All 30 samples were processed together with an estimated reagent cost of $ 6.85 per sample and a turnaround time of 24-48 hours from VTM *sample-in* to sequencing data *results-out*. Turnaround for generating sequencing data could be reduced to less than 24 hours by using very short read sequencing^12^, which however would require additional evaluation of our bioinformatics pipeline.

We were then interested to evaluate the performance of mVseq for RNA extracted from NP swab-VTM specimens. Extracted RNA has been the standard input specimen for RT-qPCR assays in the clinical laboratory^2^. We extracted RNA from NP swab-VTM specimens for eight samples with lower viral load (Ct N1, 31.6-37.2) and from one sample with lower Ct N1 of 20.1 (**Table 1**) using the Zymo Quick-DNA/RNA Viral MagBead kit (Catalogue #R2141). 100 μl of the VTM was used as input and eluted in 25 μl, of which 5 μl was used in the mRT-PCR. Using the Bioanalyzer for detection, all low viral-load samples showed at least one SARS-CoV-2 amplicon peak and the human *RPP30* peak (**Supplemental Figure 2**). Using sequencing-based detection, four of the eight low-viral load samples (S11, S13, S14, S15) were found positive for all four m-RT-PCR amplicons, three samples (S12, S17, S18) were positive for two viral amplicons and *RPP30*, while one sample (S16) was positive for only one viral amplicon and *RPP30* (**Figure 3B**). Thus, using extracted RNA from NP swab-VTM samples as the input specimen, our mVseq assay was able to correctly detect all positive samples that had an N1 Ct value of 37.2 or lower by the clinical RT-qPCR assay. The reduced sensitivity for using NP swab-VTM specimens directly without the RNA extraction step was likely due to the smaller amount of viral copies added into the m-RT-PCR. The maximum volume of NP swab-VTM specimen that could be added to the m-RT-PCR was 5.5 μl (e.g., 10.99 μl of VTM/QE mixture). For comparison in the clinical laboratory protocol, the 5 μl of extracted RNA used in testing correspond to 20 μl of NP swab-VTM specimens (a ∼4-fold increased concentration) as 200 μl of the primary NP swab-VTM specimens are eluted into 50 μl during RNA purification. In future experiments, VTM/QE mixture samples could be concentrated (f.e., using magnetic beads) to increase assay sensitivity for detecting lower viral-loads from a primary clinical specimen.

A major limitation of the m-RT-PCR-based approach is the need to open tubes for SARS-CoV-2 amplicon detection and sequencing. In comparison, viral nucleic acid testing using gold-standard RT-qPCR does not require opening tubes after amplification and thus greatly minimizes the risk for cross contamination. Additionally, many COVID-19 samples have very high viral loads (Ct value for N1 gene of 7-15) and cross contamination between samples is a major concern. To minimize contamination, we followed procedures established for working in clinical molecular laboratories including separation of pre-PCR and post-PCR laboratory space, the use of a dedicated hood for making dilutions, sterilizing Eppendorf pipettes with 10% bleach followed by UV-light treatment for 30 minutes. While a small number of background reads was seen in negative controls (<10 reads, **Figure 3A**), those could be distinguished from the multifold higher amplicon read levels in test samples. To avoid cross contamination during consecutive sequencing runs, instruments were washed with bleach according to Illumina’s guidelines before every run^12,15^.

## Conclusion

Improved molecular screening and diagnostic tools are needed to substantially increase SARS-CoV-2 testing capacity and throughput while reducing the time to receive test results. Here we developed a two-pronged approach for SARS-CoV-2 nucleic acid testing for two common detection technologies. This choice was made to provide new solutions for both rapid (using DNA electrophoresis) and high-throughput (using sequencing) readouts for SARS-CoV-2 nucleic acid testing. Notably, both readout modalities are coupled with the same multiplex RT-PCR assay, which is flexible and scalable to incorporate the larger SARS-CoV-2 genome sequence and additional RNA viruses. For RNA extraction followed by multiplex RT-PCR, all positive samples with a N1 Ct value of 37.2 or lower could be detected using either sequencing or the Bioanalyzer as readout. Eliminating the initial RNA extraction reduced the sensitivity for both modalities, and we were still able to accurately detect SARS-CoV-2 in all specimens with high viral loads. We envision the low cost and ease-of-use of the DNA electrophoresis-based readout as an attractive solution for rapid screening, and particularly in settings with constrained resources.

## Data Availability

Data is available upon request

## Acknowledgement

We thank Annie Wylie and Nathan Grubaugh for providing synthetic viral RNA fragments, and Shrikant Mane and the YCGA staff for their support and advice with this project.

## Author Contribution

NG, CS designed the assay, NG performed the experiments and developed the assay, EF and NC provided clinical samples, IT extracted and provided RNA specimens, JK established the pipeline for sequence data analysis, KB, AG, and PS provided assay reagents and advise, and NG and CS wrote the manuscript, which all authors edited and approved.

---

**Supplemental Figure 1:**
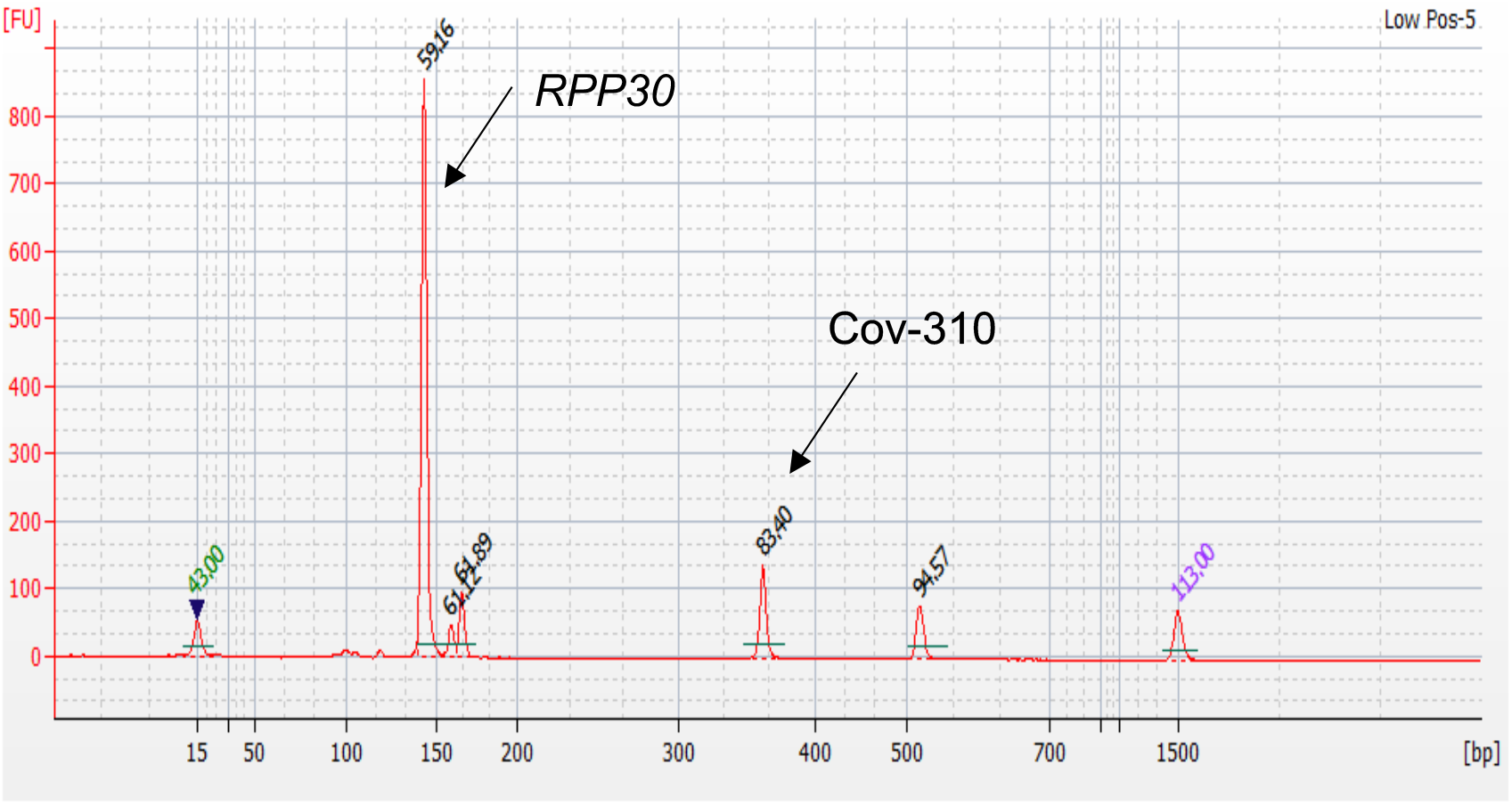
Detection of a low SARS-CoV-2 viral load sample from VTM. Agilent Bioanalyzer profile of a NP swab-VTM specimen with low viral loads (S15, Ct 31.6) shows amplification peaks for SARS-CoV-2 (Cov-310) and the human RPP30 control gene.

**Supplemental Figure 2.**
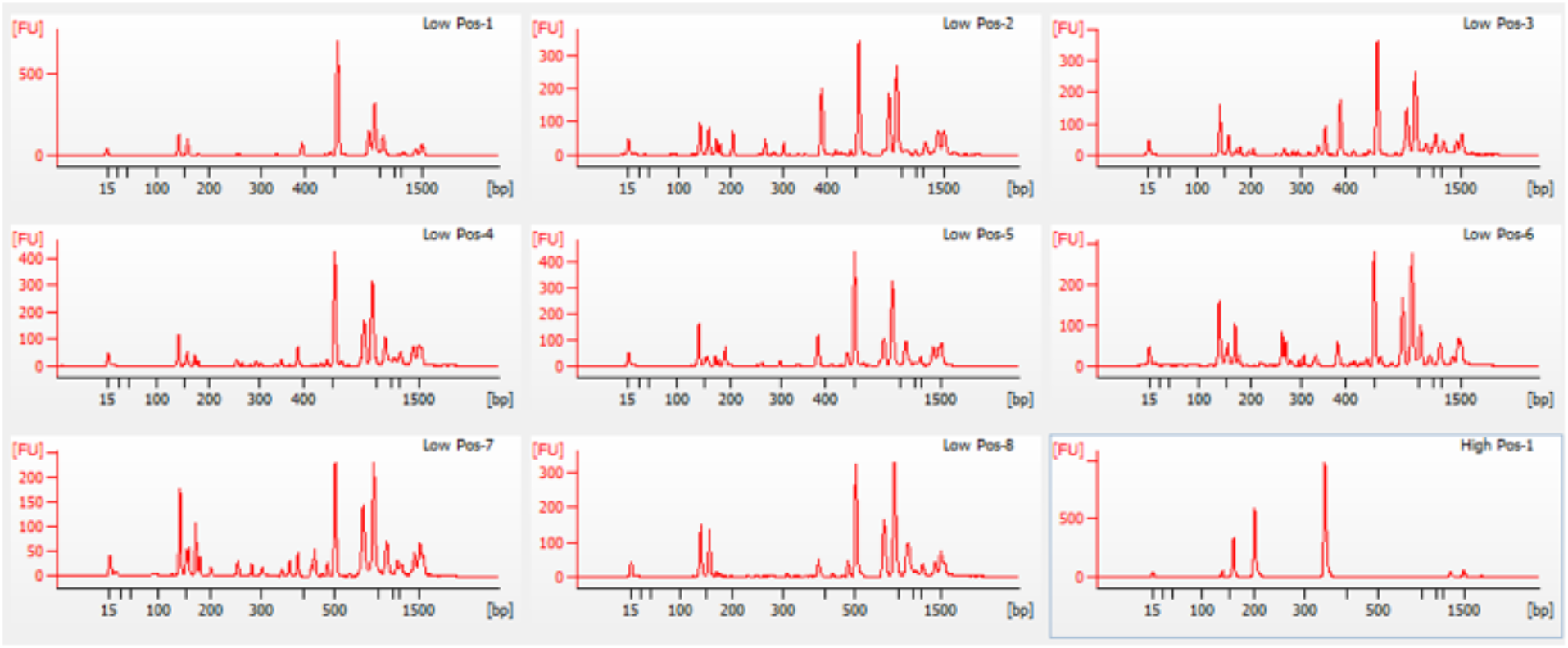
Detection of low SARS-CoV-2 viral loads from RNA specimens. Agilent Bioanalyzer profiles of RNA specimens extracted from NP swab-VTM specimens with low viral loads (31.6-37.2) show at least one amplification peak for SARS-CoV-2 and *RPP30* control gene. In comparison, the profile of the RNA specimen from a high viral load sample in panel 9 (S01, Ct 20.1) shows four peaks including the human *RPP30* amplicon and the three SARS-CoV-2 amplicons.

## References

1. CDC 2019-Novel Coronavirus (2019-nCoV) Real-Time RT-PCR Diagnostic Panel. https://www.fda.gov/media/134922/download. 2020.

2. Hanson KE, Caliendo AM, Arias CA, et al. Infectious Diseases Society of America Guidelines on the Diagnosis of COVID-19. Clin Infect Dis. 2020.

3. Williams E, Bond K, Zhang B, Putland M, Williamson DA. Saliva as a Noninvasive Specimen for Detection of SARS-CoV-2. J Clin Microbiol. 2020;58(8).

4. Pasomsub E, Watcharananan SP, Boonyawat K, et al. Saliva sample as a non-invasive specimen for the diagnosis of coronavirus disease 2019: a cross-sectional study. Clin Microbiol Infect. 2020.

5. Kojima N, Turner F, Slepnev V, et al. Self-Collected Oral Fluid and Nasal Swab Specimens Demonstrate Comparable Sensitivity to Clinician-Collected Nasopharyngeal Swab Specimens for the Detection of SARS-CoV-2. Clin Infect Dis. 2020.

6. Wyllie AL, Fournier J, Casanovas-Massana A, et al. Saliva or Nasopharyngeal Swab Specimens for Detection of SARS-CoV-2. N Engl J Med. 2020;383(13):1283–1286.

7. Landry ML, Criscuolo J, Peaper DR. Challenges in use of saliva for detection of SARS CoV-2 RNA in symptomatic outpatients. J Clin Virol. 2020;130:104567.

8. Vogels CBF, Watkins AE, Harden CA, et al. SalivaDirect: A simplified and flexible platform to enhance SARS-CoV-2 testing capacity. medRxiv. 2020:2020.2008.2003.20167791.

9. Lalli MA, Langmade SJ, Chen X, et al. Rapid and extraction-free detection of SARS-CoV-2 from saliva by colorimetric reverse-transcription loop-mediated isothermal amplification. Clin Chem. 2020.

10. Joung J, Ladha A, Saito M, et al. Detection of SARS-CoV-2 with SHERLOCK One-Pot Testing. N Engl J Med. 2020;383(15):1492–1494.

11. Fozouni P, Son S, Dí az de León Derby M, et al. Direct detection of SARS-CoV-2 using CRISPR-Cas13a and a mobile phone. medRxiv. 2020:2020.2009.2028.20201947.

12. Bloom JS, Jones EM, Gasperini M, et al. Swab-Seq: A high-throughput platform for massively scaled up SARS-CoV-2 testing. medRxiv. 2020.

13. Gorzynski JE, De Jong HN, Amar D, et al. High-throughput SARS-CoV-2 and host genome sequencing from single nasopharyngeal swabs. medRxiv. 2020.

14. Huang J, Zhao L. A high-throughput strategy for COVID-19 testing based on next-generation sequencing. medRxiv. 2020:2020.2006.2012.20129718.

15. Yelagandula R, Bykov A, Vogt A, et al. SARSeq, a robust and highly multiplexed NGS assay for parallel detection of SARS-CoV2 and other respiratory infections. medRxiv. 2020:2020.2010.2028.20217778.

16. Tan SK, Shen P, Lefterova MI, et al. Transplant Virus Detection Using Multiplex Targeted Sequencing. J Appl Lab Med. 2018;2(5):757–769.

17. Ladha A, Joung J, Abudayyeh O, Gootenberg J, Zhang F. A 5-min RNA preparation method for COVID-19 detection with RT-qPCR. medRxiv. 2020:2020.2005.2007.20055947.

18. Shirato K, Nao N, Katano H, et al. Development of Genetic Diagnostic Methods for Detection for Novel Coronavirus 2019(nCoV-2019) in Japan. Jpn J Infect Dis. 2020;73(4):304–307.

